# Assessing the impact of a national clinical guideline for the management of chronic pain on opioid prescribing rates: A controlled interrupted time series analysis

**DOI:** 10.1101/2021.02.19.21251770

**Authors:** Harry L. Hébert, Daniel R. Morales, Nicola Torrance, Blair H. Smith, Lesley A. Colvin

## Abstract

**Background:** Opioids can be effective analgesics, but long-term use may be associated with harms. In 2013, the first national, comprehensive, evidence-based pain management guideline was published, from the Scottish Intercollegiate Guideline Network (SIGN 136: Management of Chronic Pain) with key recommendations on analgesic prescribing. This study aimed to examine the potential impact on national opioid prescribing rates in Scotland.

**Methods:** Trends in national and regional community opioid prescribing data for Scotland were analysed from quarter one (Q1) 2005 to Q2 2020. Interrupted time series regression examined the association of SIGN 136 publication with prescribing rates for opioid-containing drugs. Gabapentinoid prescribing was used as an outcome control.

**Results:** After a positive prescribing trend pre-publication, the timing of SIGN 136 publication was associated with a negative change in trend of opioid prescribing rates (−2.82 items per 1,000 population per quarter [PTPPQ]; P<0.01). By Q2 2020, the relative reduction in opioid prescribing rate was -20.67% (95% CI: -23.67, -17.77). This persisted after controlling for gabapentinoid prescribing and was mainly driven by reduction in weak opioids, whereas strong opioid prescribing rates continued to rise. Gabapentinoid prescribing showed a significant rise in level (8.00 items per 1,000 population; P=0.01) and trend (0.27 items PTPPQ; P=0.01) following SIGN 136 publication.

**Conclusions:** Publication of SIGN 136 was associated with a reduction in opioid prescribing rates. This suggests that changes in clinical policy through evidence-based national clinical guidelines may affect community opioid prescribing, though this may be partially replaced by gabapentinoids, and other factors may also contribute.

## Introduction

Chronic pain is a common and complex problem, with a debilitating impact on quality of life^1^. Whilst there is often no cure, opioids have been commonly used to treat patients with the disorder^2^. There is good evidence for their efficacy in acute and cancer-related pain, but very limited high-quality evidence for effectiveness in long-term management of chronic non-malignant pain^3^. Inadequate pain relief with opioids can lead to dose escalation and tolerance, with risks of major adverse events such as dependence, addiction, overdose and death^4^.

Opioid use has been increasing steadily worldwide, with the World Drug Report – 2021 estimating that nearly 62 million people (aged 15-64) used opioids globally in 2019^5^. In particular, the increase in prescribing of opioids in the USA from the late 1990s to the early 2010s has been well documented^6^. Possible explanations for this increase include an ageing population at greater risk of developing chronic pain conditions, the publication of clinical guidelines recommending opioids for chronic non-cancer pain, despite insufficient good quality evidence, (non-evidence-based) changes in recommendations on use from professional bodies, and effective marketing from pharmaceutical companies^7^. This increase in opioid use has given rise to the “opioid epidemic” in the USA and there are concerns a similar situation could be happening in the UK, including Scotland^8 9^.

In December 2013, the Scottish Intercollegiate Guideline Network (SIGN), in consultation with the National Chronic Pain Steering Group of the Scottish Government and key stakeholders, published the first comprehensive evidence-based guideline for the assessment and management of chronic, non-malignant pain in adults (SIGN 136)^10^. SIGN 136 identified a research gap around understanding rates and effects of opioid prescribing in Scotland^11^. A resulting investigation of Scottish data revealed that prescribing rates of strong opioids doubled in the 10-year period leading up to publication of SIGN 136 (2003-2012)^12^. One aspiration of SIGN 136 was to influence safe and appropriate use of opioids in chronic pain, reducing unnecessary and potentially harmful prescribing. Whilst it is recognised that clinical practice guidelines have the potential to increase the quality of care, systematic analysis of their impact is not common^13^.

Gabapentinoids (mainly gabapentin and pregabalin) are licensed for the treatment of peripheral and central neuropathic pain, with strong evidence for their effectiveness^14 15^. In contrast to opioids, SIGN 136 key recommendations did not caution about risks of harm related to gabapentinoid misuse, as, at the time of publication, there was little evidence of this.

Opioid prescribing rates have now become one of National Health Service (NHS) Scotland’s key National Therapeutic indicators, a set of prescribing indicators in specific therapeutic areas that can be used to compare prescribing behaviours against established guidelines^16^. These indicators clearly show that opioid prescribing rates are beginning to fall across Scotland. However, to the best of our knowledge, the potential impact of SIGN 136 on opioid prescribing rates has not been investigated. Therefore, this study aimed to analyse opioid prescribing rates in Scotland before and after SIGN 136 publication and to compare these to gabapentinoids.

## Methods

We followed the Framework for Enhanced Reporting of Interrupted Time Series Studies (FERITS) statement^17^, an adaptation of the Transparent Reporting of Evaluations with Nonrandomised Designs (TREND) statement^18^, for the reporting of this study (Supplementary Table S1).

### Study Design

This study was an interrupted time series analysis of national level prescribing data on opioid analgesics prescribed in primary care and dispensed in by community pharmacies, to test the hypothesis that SIGN 136 is associated with a significant change in opioid prescribing trend. Gabapentinoid prescribing data were also obtained and used to control for potential confounding from other interventions. The study period was from January 2005 to June 2020 and incorporated the SIGN 136 publication date (December 2013).

### Data Source

Data on all opioids and gabapentinoids, prescribed through primary care (in the community) by general practitioners (GPs) and non-medical prescribers and dispensed by community pharmacists in Scotland, were obtained from Public Health Scotland (PHS). This was based on aggregated and publicly available data^19^. PHS (https://www.publichealthscotland.scot/) is part of NHS Scotland and holds individual-level prescribing data through the Prescribing Information System (PIS), which is a national data system, set up in 2009, that captures all NHS prescriptions dispensed and reimbursed in the community (https://www.ndc.scot.nhs.uk/National-Datasets/data.asp?SubID=9). In the UK, healthcare policy is devolved to the individual nations and, in Scotland, community prescriptions are free at the point of delivery. Pharmacists are reimbursed for the prescriptions they dispense. The PIS covers the population of Scotland (approximately 5.3 million), with GPs accounting for approximately 95% of community prescribing, and a capture rate of 98.7% from prescription forms^20^. Also included in the data request were annual mid-year population estimates for Scotland as of 30^th^ June each year.

Formal ethical approval was not required as the study used publicly available data which contained no patient or prescriber identifiable information.

### Publication of SIGN 136

The Scottish Intercollegiate Guideline Network (SIGN) was established in 1993 by the Scottish Medical Royal Colleges and is now part of Healthcare Improvement Scotland, part of NHS Scotland. It produces evidence-based clinical practice guidelines for use across NHS Scotland, with accredited methodology^21^, and is a member organisation of the Guidelines International Network (https://g-i-n.net/). SIGN 136 was published in December 2013 and, after a systematic review of the evidence, included key recommendations and best practice statements on safe and effective opioid prescribing (Supplementary Box S1). The Scottish Government requires Health Boards to identify areas of concern where they are not meeting SIGN’s Key recommendations, so they become important benchmark standards for care. The Scottish Government also provides regular feedback of opioid prescribing data to individual GP Practices as National Therapeutic Indicators, to ensure implementation of SIGN 136. Therefore, it has formed the basis of pain service provision and improvement in Scotland since its publication. The guideline is aimed at all healthcare professionals involved in the assessment and management of adult patients with chronic non-malignant pain in non-specialist settings. At the time of publication, hard copies were disseminated to all primary care practices across Scotland and the guideline is available for download from SIGN’s website (https://www.sign.ac.uk/assets/sign136.pdf). A patient version was also available^22^.

The opioids section of this guideline (section 5.3 “Opioids”) was subsequently updated in August 2019^23^. However, for this study we only considered the original 2013 publication as the “intervention”.

To control for any potential unforeseen confounders acting in Scotland, such as changes in prevalence of chronic pain or changes in policy involving the use of pharmacological interventions for chronic pain in general, we decided to use gabapentinoid prescribing as a comparison. SIGN 136 included guidance for gabapentinoids (gabapentin and pregabalin), though in contrast to opioids there were no specific recommendations warning of the potential for abuse, addiction or other side effects (Supplementary Box S2). Therefore, it was hypothesised that gabapentinoid prescribing rates would not have reduced as a result of SIGN 136 publication.

### Outcome

The primary outcome was the number of opioid prescription items dispensed per 1,000 population per quarter (PTPPQ). A prescription may contain multiple pharmaceutical products. If a prescription form includes three medicines, it is counted as three prescription items. The control outcome was the number of gabapentinoid (gabapentin and pregabalin) items dispensed PTPPQ. Quarters were defined as January to March (Q1), April to June (Q2), July to September (Q3) and October to December (Q4), inclusive. A list of all relevant opioid-containing drugs included in the study is given in Supplementary Table S2. These include single and compound analgesics found in chapter 4.7.2 (“Opioid analgesics”) of the British National Formulary (BNF)^24^ as well as additional combination products of opioids (e.g. co-codamol) found elsewhere. Gabapentin and pregabalin are detailed in chapter 4.8.1 (“Control of epilepsy”) of the BNF. The dataset includes all items prescribed through the NHS in primary care in Scotland (which provides the first point of contact for patients in the healthcare system, usually through GP practices), dispensed in the community in the UK and submitted for reimbursement. Data on items prescribed but not subsequently submitted for dispensing by the patient (estimated to be ∼6%)^20^ or dispensed but not submitted for reimbursement by the pharmacy are not currently held by PHS. The small number of private prescriptions, hospital and direct supply of medicines to patients (e.g. prescriptions supplied through specialist clinics) were not included.

### Statistical Analysis

Linear regression was used to analyse the impact of SIGN 136 on opioid and gabapentinoid prescribing trends nationally. The model for the analysis is provided in Box 1. As part of this process, plots of each time series were studied to check that the assumption of linearity was appropriate in each analysis.

The analysis of opioid prescribing trends was stratified according to opioid strength (weak or strong), and recipients’ age (0-29, 30-49, 50-69 and 70+ years) and gender. The stratification of drugs by strength was based on their status in SIGN 136 with codeine, dihydrocodeine, meptazinol and tramadol considered weak opioids and buprenorphine, diamorphine, dipipanone, fentanyl, hydromorphone, methadone, morphine, oxycodone, papaveretum, pentazocine, pethidine and tapentadol considered strong opioids. Compound drugs were classified according to the parent opioid (Supplementary Table S2). Level and slope change models were used to test the hypothesised immediate and longer-term impact on prescribing behaviour that the publication of SIGN 136 would have had.

A controlled interrupted time series approach was used to compare opioid prescribing trends to gabapentinoid prescribing trends; weak opioid vs strong opioid prescribing; and prescribing between men and women.

All analyses used data from Q1 2005 to Q2 2020 inclusive, apart from the analyses involving age and gender, where data were only available between Q1 2010 and Q2 2020. The publication of SIGN 136 (i.e. the “intervention”) was defined as Q4, 2013, providing 36 data points before the publication (16 data points for the age and gender analyses) and 26 data points after the publication.

The effect of the publication was presented as the relative percentage change in prescribing rate at Q2 2020 compared to the predicted value at the same time point had pre-publication trends continued (the counterfactual, calculated from the model coefficient estimates). The 95% confidence intervals (CI) were calculated using model bootstrapping approaches^25^. All models were checked for autocorrelation using the Durbin-Watson statistic. A range of 1.50-2.50 was used to indicate an acceptable level of autocorrelation. Models outside this range were corrected using Newey-West standard errors.

All analyses were carried out using the statistical software R (version 4.0.3)^26^.

## Results

### Summary Statistics

The mid-year population estimates from 2005 to 2020 for the whole of Scotland are given in Supplementary Table S3. The estimated population size of Scotland was 5,110,200 in 2005 and 5,466,000 in 2020. A breakdown of the mid-year population estimates by age and gender is given in Supplementary Table S4 from 2010-2020.

Between Q1 2005 and Q2 2020, a total of 91,210,542 prescription items of the specified opioid or opioid-containing combination drugs included in this study were dispensed across Scotland. At the same time, a total of 12,036,499 prescriptions items of gabapentinoids were dispensed across Scotland. The total number of items of each drug prescribed is given in Supplementary Table S2.

### Overall Analysis

Across the whole of Scotland, the number of opioid prescription items rose from 1,040,276 in Q1 2005 to 1,608,984 in Q4 2013, an increase of 54.7%. Since the publication of SIGN 136 (Q4 2013), the number of opioid prescriptions has gradually fallen to 1,499,400 items in Q2 2020, a decrease of 6.8% (Figure 1).

**Figure 1.**
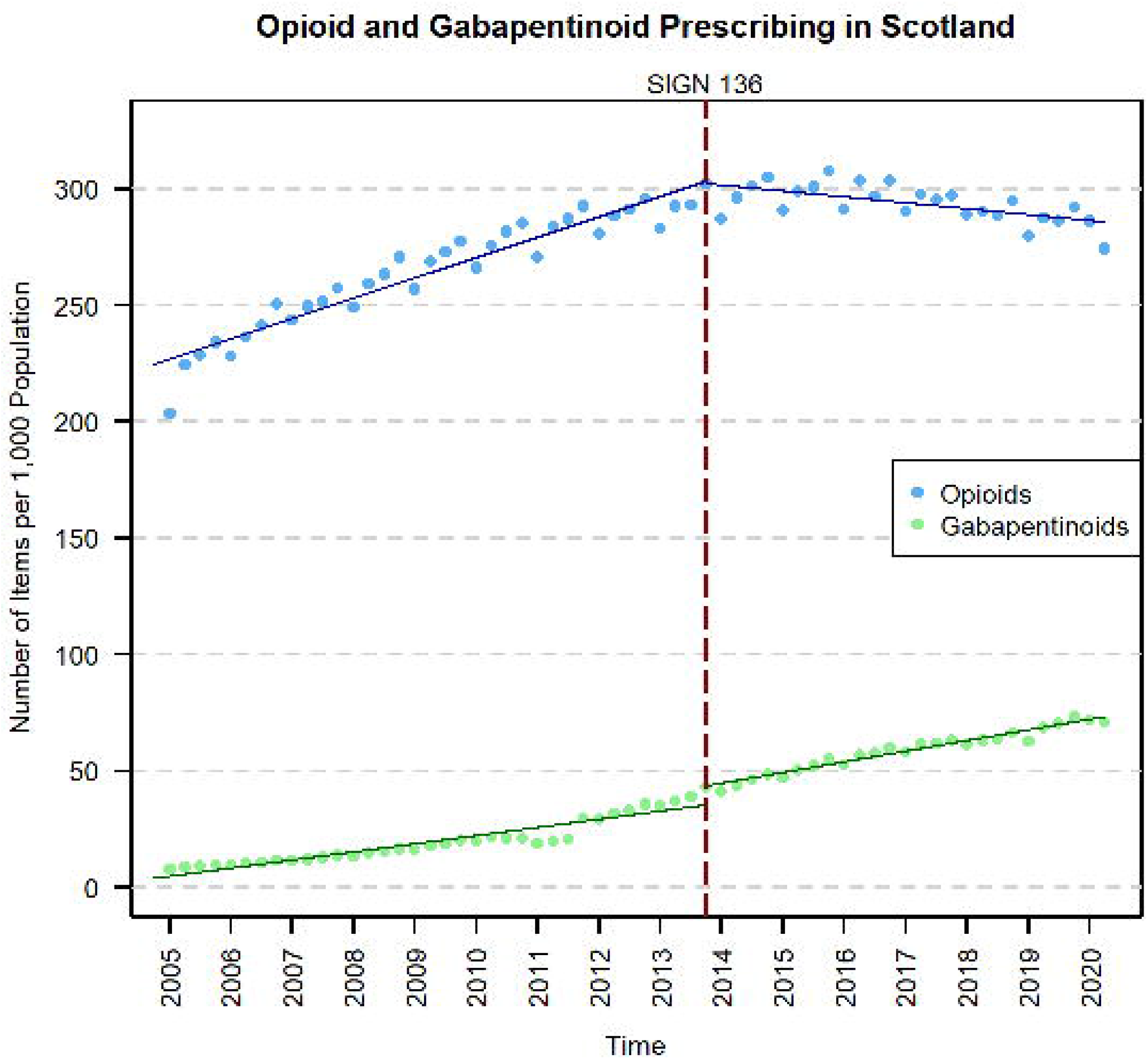
Opioid prescribing time series in Scotland before and after the publication of SIGN 136 in December 2013 (red dashed line). The solid lines represent the prescribing trend derived from the interrupted time series analysis.

There was a significant positive trend in opioid prescribing rates pre-publication (2.19 items PTPPQ; P<0.01), followed by no significant level change, and a significant change in trend following the publication (−2.82 items PTPPQ; P<0.01). Opioid prescribing rates began to fall post-publication at -0.64 items PTPPQ and at the end of the study period the relative change was estimated to be -20.67% (95% CI: -23.67, - 17.77) compared to the counterfactual (Table 1).

**Table 1.** Single-group interrupted time series analysis of opioid and gabapentinoid prescribing in Scotland

|  | Opioids |  |  | Gabapentinoids |  |  |
| --- | --- | --- | --- | --- | --- | --- |
|  | Estimate<br>(95% Confidence Interval) | Standard Error | P value | Estimate<br>(95% Confidence Interval) | Standard Error<sup>a</sup> | P value |
| Intercept, $\beta_0$ | 224.46 (219.79, 229.12) | 2.33 | <0.01 | 3.62 (0.35, 6.90) | 1.64 | 0.03 |
| Pre-Intervention Trend, $\beta_1$ | 2.19 (1.97, 2.41) | 0.11 | <0.01 | 0.88 (0.67, 1.08) | 0.10 | <0.01 |
| Change in Level, $\beta_2$ | -1.09 (-8.21, 6.03) | 3.56 | 0.76 | **8.00 (2.27, 13.73)** | **2.86** | **0.01** |
| **Change in Trend, $\beta_3$** | **-2.82 (-3.24, -2.40)** | **0.21** | **<0.01** | **0.27 (0.07, 0.47)** | **0.10** | **0.01** |
| Post-Intervention Trend, $\beta_{1+3}$ | -0.64 (-0.98, -0.29) | 0.17 | <0.01 | 1.15 (1.04, 1.26) | 0.05 | <0.01 |
| **Relative Change, %<sup>b</sup>** | **-20.67 (-23.67, -17.77)** | **n/a** | **n/a** | **25.89 (15.13, 42.84)** | **n/a** | **n/a** |
| Durbin-Watson Statistic | 1.64 | n/a | n/a | 0.80 | n/a | n/a |

$\beta_{0-3}$ are coefficients from the single-group interrupted time series analysis model; the intercept represents the outcome at the start of the study period; the relative change is calculated compared to the predicted value at the same time point had the pre-intervention trend continued.
<sup>a</sup>Newey-West
<sup>b</sup>Calculated at quarter 2 2020. 95% confidence interval calculated using model bootstrapping.

In comparison, there was a significant positive trend in gabapentinoid prescribing pre-publication (0.88 items PTPPQ; P<0.01), followed by a significant increase in level (8.00 items per 1,000 population [PTP]; P<0.01), and a significant positive change in trend post-publication (0.27 items PTPPQ; P=0.01). The interrupted time series analysis and prescribing rates for gabapentinoids are presented in Table 1 and Figure 1.

When opioid prescribing was adjusted for gabapentinoid prescribing (Table 2), the significant change in trend post-publication was maintained (−3.09 items PTPPQ; P<0.01). There was also a significant negative difference in level change compared to gabapentinoids (−9.09 items PTP; P=0.02). The adjusted publication effect on opioid prescribing rate at the end of the study period was calculated to be -24.85% (95% CI: - 28.25, -21.61).

**Table 2.**
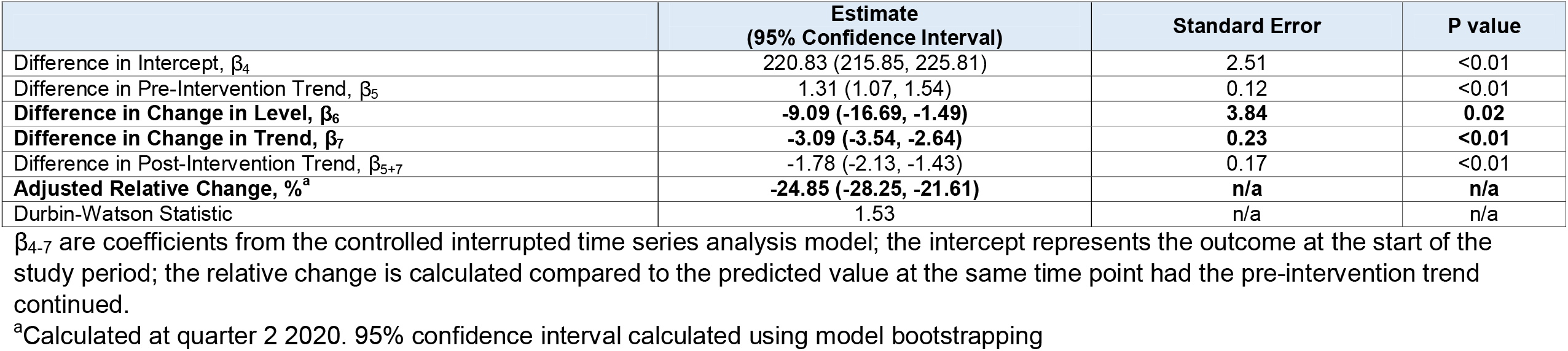
Results of the controlled interrupted time series analysis of opioid prescribing rates adjusted for gabapentinoid prescribing in Scotland

|  | Estimate<br>(95% Confidence Interval) | Standard Error | P value |
| --- | --- | --- | --- |
| Difference in Intercept, $\beta_4$ | 220.83 (215.85, 225.81) | 2.51 | <0.01 |
| Difference in Pre-Intervention Trend, $\beta_5$ | 1.31 (1.07, 1.54) | 0.12 | <0.01 |
| **Difference in Change in Level, $\beta_6$** | **-9.09 (-16.69, -1.49)** | **3.84** | **0.02** |
| **Difference in Change in Trend, $\beta_7$** | **-3.09 (-3.54, -2.64)** | **0.23** | **<0.01** |
| Difference in Post-Intervention Trend, $\beta_{5+7}$ | -1.78 (-2.13, -1.43) | 0.17 | <0.01 |
| **Adjusted Relative Change, %<sup>a</sup>** | **-24.85 (-28.25, -21.61)** | **n/a** | **n/a** |
| Durbin-Watson Statistic | 1.53 | n/a | n/a |

$\beta_{4-7}$ are coefficients from the controlled interrupted time series analysis model; the intercept represents the outcome at the start of the study period; the relative change is calculated compared to the predicted value at the same time point had the pre-intervention trend continued.
<sup>a</sup>Calculated at quarter 2 2020. 95% confidence interval calculated using model bootstrapping

### Stratified Analysis

When stratifying opioid prescribing by strength (Figure 2), both weak (−2.27 items PTPPQ; P<0.01) and strong opioids (−0.55 items PTPPQ; P<0.01) showed significant negative changes in trend post-publication, but non-significant changes in level (Table 3). However, there was a significantly greater negative change in trend for weak opioids than for strong opioids in the adjusted analysis (−1.72 items PTPPQ; P<0.01; Supplementary Table S5). Post-publication, weak opioid prescribing rates began to fall at -0.91 items PTPPQ, whereas strong opioids rates continued to rise at 0.27 items PTPPQ. The relative change was estimated to be -21.68% (95% CI: -24.75, -18.57) for weak opioids and -17.49% (95% CI: -20.15, -14.87) for strong opioids.

**Table 3.**
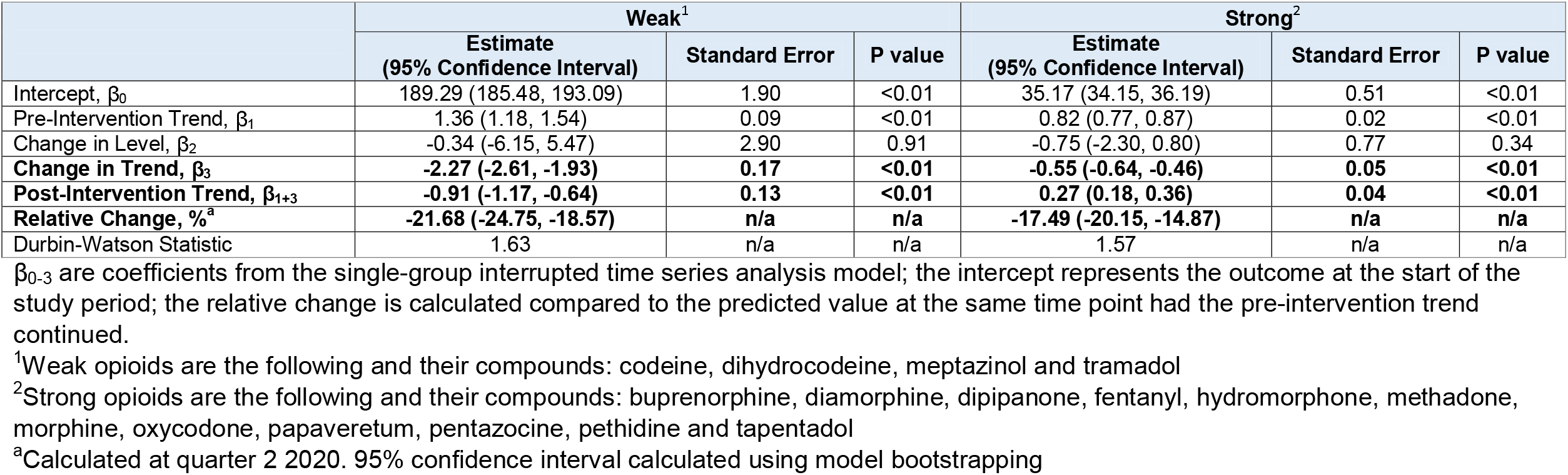
Results of the single interrupted time series analyses of weak and strong opioid prescribing rates in Scotland

|  | Weak<sup>1</sup> |  |  | Strong<sup>2</sup> |  |  |
| --- | --- | --- | --- | --- | --- | --- |
|  | Estimate<br>(95% Confidence Interval) | Standard Error | P value | Estimate<br>(95% Confidence Interval) | Standard Error | P value |
| Intercept, $\beta_0$ | 189.29 (185.48, 193.09) | 1.90 | <0.01 | 35.17 (34.15, 36.19) | 0.51 | <0.01 |
| Pre-Intervention Trend, $\beta_1$ | 1.36 (1.18, 1.54) | 0.09 | <0.01 | 0.82 (0.77, 0.87) | 0.02 | <0.01 |
| Change in Level, $\beta_2$ | -0.34 (-6.15, 5.47) | 2.90 | 0.91 | -0.75 (-2.30, 0.80) | 0.77 | 0.34 |
| **Change in Trend, $\beta_3$** | **-2.27 (-2.61, -1.93)** | **0.17** | **<0.01** | **-0.55 (-0.64, -0.46)** | **0.05** | **<0.01** |
| **Post-Intervention Trend, $\beta_{1+3}$** | **-0.91 (-1.17, -0.64)** | **0.13** | **<0.01** | **0.27 (0.18, 0.36)** | **0.04** | **<0.01** |
| **Relative Change, %<sup>a</sup>** | **-21.68 (-24.75, -18.57)** | **n/a** | **n/a** | **-17.49 (-20.15, -14.87)** | **n/a** | **n/a** |
| Durbin-Watson Statistic | 1.63 | n/a | n/a | 1.57 | n/a | n/a |

$\beta_{0-3}$ are coefficients from the single-group interrupted time series analysis model; the intercept represents the outcome at the start of the study period; the relative change is calculated compared to the predicted value at the same time point had the pre-intervention trend continued.
<sup>a</sup>Calculated at quarter 2 2020. 95% confidence interval calculated using model bootstrapping

**Figure 2.**
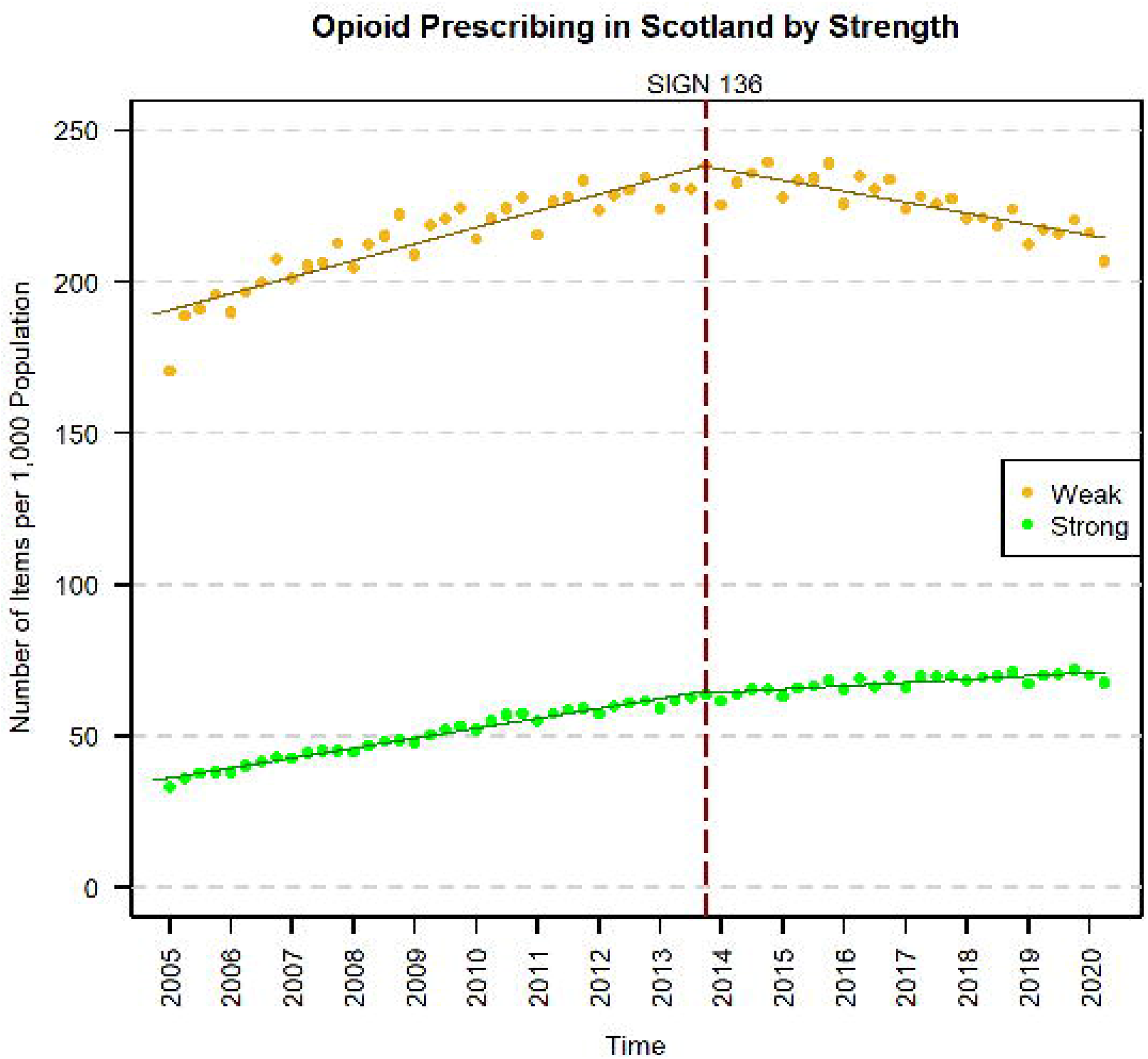
Prescribing time series of weak^1^ and strong^2^ opioids in Scotland, before and after the publication of SIGN 136 in December 2013 (red dashed line). The solid lines represent the prescribing trend derived from the interrupted time series analysis. ^1^Weak opioids are the following and their compounds: codeine, dihydrocodeine, meptazinol and tramadol ^2^Strong opioids are the following and their compounds: buprenorphine, diamorphine, dipipanone, fentanyl, hydromorphone, methadone, morphine, oxycodone, papaveretum, pentazocine, pethidine and tapentadol

In the gender analysis for opioids, both women (−3.26 items PTPPQ; P<0.01) and men (−2.52 items PTPPQ; P<0.01) showed significant negative changes in trend post-publication, but non-significant changes in level (Supplementary Figure S1). Opioid prescribing rates began to fall post-publication for both women (−0.54 items PTPPQ) and men (−0.44 items PTPPQ; Supplementary Table S6). There were no significant differences between the genders in the adjusted analysis, in terms of the post-publication effects (Supplementary Table S7) and the relative change was estimated to be -19.11% (95% CI: -23.61, -13.13) for women and -21.54% (95% CI: -25.88, - 15.64) for men.

Finally, in the age analysis (Supplementary Figure S2) all groups showed non-significant changes in level except the >70 years old group (13.61 items PTP; P=0.03) and all the groups showed significant negative changes in trend post-publication (0-29 years: -0.74; 30-49 years: -5.47; 50-69 years: -3.21; >70 years: -3.46 items PTPPQ respectively). All the age groups showed significant negative post-publication trends (0-29 years: -0.41; 30-49 years: -1.15; >70 years: -0.27 items PTPPQ respectively), except the 50-69 year group. The relative changes were estimated to be -36.13 (95% CI: -44.43, -22.56) in 0-29 years, -31.64 (95% CI: -35.43, -26.81) in 30-49 years, - 15.29 (95% CI: -21.33, -6.24) in 50-69 years and -12.04 (95% CI: -16.25, -6.33) in >70 years (Supplementary Table S8).

## Discussion

### Summary

In this study, publication of a national clinical guideline on management of chronic pain (SIGN 136) in 2013 was associated with a significant negative change in trend in primary care opioid prescribing in Scotland that resulted in a relative reduction of 21% by Q2 2020. This finding persisted when controlling for gabapentinoids, which was not associated with any similar changes in prescribing trend. Stratified analyses by opioid strength, age category and gender showed that SIGN 136 was associated with a significant negative change in trend in all groups. Despite this, prescribing rates of strong opioids continued to rise post-publication, albeit at a slower rate than pre-publication. To the best of our knowledge this is the first study to analyse changes in opioid prescribing trends in Scottish primary care, and to examine the association of prescribing rates with a specific intervention intended to influence these.

### Interpretation

Increasing opioid prescribing in the UK and elsewhere has been well documented^27–30^. This is also the case in Scotland where prescribing of opioids has increased both locally and nationally^9 12^. However, the time-period covered by the Scottish studies was prior to the publication of SIGN 136. Similar increases have been identified more recently across the UK^31^. This continuing increase in opioid prescribing rate across the UK beyond 2013 appears to be in contrast with the results from the current study.

However, we found that this decrease in prescribing numbers and most of the change in trend associated with SIGN 136 was being driven by weak opioids, with strong opioid prescribing rates continuing to increase. This increase in strong opioid prescribing rates appears to be in line with a previous study in Wales^27^. This may reflect a marked change in prescribing behaviour for weak opioids, whilst changes in prescribing behaviour for strong opioids has been slower. However, weak opioids continued to be much more frequently prescribed than strong opioids overall.

In addition to the opioid findings, there was also a significant increase in gabapentinoid prescribing trend. A possible reason for this could be an increase in the number of neuropathic pain cases being diagnosed and treated, particularly as gabapentinoids are recommended first or second line treatment for neuropathic pain in national and international guidelines^32 33^. However, this is unlikely to account for all of the increase in gabapentinoid prescribing rates^34^. Another potential explanation is that the publication of SIGN 136 has led to a swap of prescription of opioids for gabapentinoids. Gabapentinoids are licensed for the treatment of neuropathic pain, yet there is increasing evidence that they are being prescribed off-label for other forms of pain^35^, despite limited evidence for their effectiveness for non-neuropathic pain^36 37^. However, gabapentinoids have themselves recently been associated with increased rates of adverse outcomes and the increase in their use is a cause for concern^38 39^. This increase has prompted the reclassification of gabapentin and pregabalin as Class C controlled drugs in the UK (placing greater legal restrictions on their supply) and the complete removal of pregabalin from the formulary for neuropathic pain in Northern Ireland^40^.

### Strengths and Weaknesses

Due to the epidemiological design of the study, the impact of other guidelines, policies and related interventions within Scotland cannot be ruled out. In the UK these include the reclassification of tramadol in 2013^41^, chronic pain initatives^42^ and an online resource, “Opioids Aware”, to support prescribing of opioids for chronic pain^43^. Furthermore, initiatives outside of the UK such as the Helping to End Addiction Over the Long-term (HEAL) initiative in the USA^44^ may have further influenced more recent prescribing practices in Scotland. Media coverage of the opioid epidemic in the USA is also likely to have raised awareness amongst the general public and patients may have gained a greater understanding of the risks of opioid treatments and other options available.

In contrast to the findings in this study of decreasing opioid prescribing rates since December 2013, it is interesting to note a previous study in which regulatory warnings about the cardiovascular safety of diclofenac in Scotland appeared to increase the rate of switching to opioids around the same time as SIGN 136 publication^45^. As a result, it is possible that without this influence, the negative change in opioid prescribing trend associated with SIGN 136 may have been greater.

A revised version of SIGN 136 was also published in August 2019, providing more up to date evidence-based guidance on opioid prescribing^23^. Through both this and the original version, SIGN 136 has been influential in driving UK and Scottish Government policy on chronic pain management and has been incorporated into clinical practice with the publication of the Scottish National Prescribing Strategy for Chronic Pain^46^, the Royal College of Anaesthetists Quality Improvement Compendium^47^ and the Medicines and Healthcare products Regulatory Agency guidance on the safe use of opioids^48^. The potential impact of the update has not yet been assessed and should be the focus of future studies. So too should the potential impact of the COVID-19 pandemic, which has had a major impact on emergency and specialist care services with concern around a potential increase in opioid prescribing rates as patients turn to primary care^49^.

Regardless of the potential cause, the reduction in opioid prescribing trend described in this study demonstrates the important role that evidence-based clinical guidelines potentially play in prescribing behaviours. Previous studies have indicated that GPs’ beliefs about whether or not opioids are appropriate for chronic non-cancer pain are a driver in whether to prescribe them, and a lack of a consistent approach and effective alternatives are barriers to deprescribing opioids, despite clear concerns about potential harms such as addiction, dependence and misuse^50–52^. This supports the need for dedicated guidelines, based on strong evidence.

This study also highlights interrupted time series analysis as a potential tool for assessing the impact of clinical guidelines. A previous study that used a similar approach found that the reclassification of tramadol as a Schedule 3 controlled substance in June 2014 was significantly associated with a reduction in tramadol utilisation in England and Wales^41^.

Despite focussing on prescribing rates in relation to SIGN 136, we were unable to assess other opioid-related outcomes, such as abuse, misuse and overdose. Since a key aim of SIGN 136 is to improve patient quality of life, it would be interesting to see if incidence rates for these outcomes have fallen in line with opioid prescribing since publication of SIGN 136, as would be expected given their close association^53^. Recent data from Scotland show opioid-related deaths rates are continuing to rise, though this could be due to illicit use as well as iatrogenic use^54^.

## Conclusion

In conclusion, opioid prescribing rates have been falling in Scotland since 2013. Whilst this effect cannot be definitively linked to the publication of SIGN 136, it at least suggests that changes in Scottish clinical and government policy relating to chronic pain management, most of which have been inspired by its publication, are having a positive effect on opioid prescribing practices in primary care. This highlights the importance of providing continued robust clinical advice, based on up-to-date evidence, for safe and effective treatment for chronic pain.

## Data Availability

Summary data used in this study are available from the corresponding author, upon reasonable request. Requests for community prescribing data in Scotland can be made to Public Health Scotland.

## Details of Author Contributions

Study conception and design: B.H.S. and L.A.C.

Data acquisition: H.L.H.

Data analysis and interpretation: all authors

Drafting the article and revising for important intellectual content: all authors

Final approval of the published version: all authors.

## Acknowledgements

We acknowledge the help and support of Public Health Scotland and the Health Informatics Centre, University of Dundee for managing and supplying the anonymised data.

## Conflict of Interest

L.A.C. chaired the Guideline Development Group for the Scottish Intercollegiate Guideline Network publication, “Management of Chronic Pain” to which this paper refers, proposed an update due to changing evidence, and contributed to the update of the opioids section. She has been Vice Chair of SIGN since October 2020 and was a member of the Scottish Government group that developed the National Prescribing Strategy for Chronic Pain.

B.H.S. was a member of the Guideline Development Group for the Scottish Intercollegiate Guideline Network publication, “Management of Chronic Pain” to which this paper refers and contributed to the update of the opioids section. He was the Scottish Government’s National Lead Clinician for Chronic Pain (2014 to 2021) and was a member of the Scottish Government group that developed the National Prescribing Strategy for Chronic Pain.

D.R.M. is supported by a Wellcome Trust Clinical Research Development Fellowship (Grant 214588/Z/18/Z) and reports grants from the Chief Scientist Office (CSO), Health Data Research UK (HDR-UK) and the National Institute of Health Research (NIHR), outside of the submitted work.

H.L.H., and N.T. declare that they have no conflicts of interest.

## Box 1.

Model for a controlled interrupted time series analysis

Y = β_0_ + β_1_ * Time + β_2_ * Intervention status + β_3_ * Intervention status * Time + β_4_ * Cohort status + β_5_ * Cohort status * Time + β_6_ * Cohort status * Intervention status + β_7_ * Cohort status * Intervention status * Time

Where Y is the outcome (prescribing rate).

β_0-3_ are coefficients representing the control cohort (e.g. gabapentinoid series) where β_0_ is the intercept or value of the outcome at the start of the study period, β_1_ is the change in outcome per unit time (trend) before the intervention, β_2_ is the immediate step change in level following the intervention and β_3_ is the change in trend following the intervention (relative to the trend before the intervention – β_1_). β_1_ and β_3_ can therefore be summed to provide the trend following the intervention. β_0-3_ can also be used in isolation as a standalone model for a single interrupted time series analysis.

β_4-7_ are coefficients representing the difference between the case series (e.g. opioid series) and the control series. β_4_ is the difference in intercept level, β_5_ is the difference in trend before the intervention, β_6_ is the difference in immediate change in level following the intervention and β_7_ is the difference in change in trend following the intervention. β_5_ and β_7_ can be summed to provide the difference in trend following the intervention.

Time, intervention status and cohort status relate to variables in the dataset representing the time elapsed since the start of the study period, the pre- or post-intervention period and the time series assignment (e.g. opioids or gabapentinoids).

