## Supplementary for "Assessing the impact of a national clinical guideline for the management of chronic pain on opioid prescribing rates: A controlled interrupted time series analysis"

### **Supplementary Box S1.** Summary of the original SIGN 136 guideline (2013 edition) intervention for use of opioids in the management of adults with chronic non-malignant pain in non-specialist settings

**Key recommendation(s)** (based on evidence)

- Strong opioids should be considered as an option for pain relief for patients with chronic low back pain or osteoarthritis, and only continued if there is ongoing pain relief. Regular review is required.

**Other recommendation(s)** (based on evidence)

- Patients prescribed opioids should be advised of the likelihood of common side effects such as nausea and constipation.
- It may be necessary to trial more than one opioid sequentially, as both effectiveness and side effects vary between opioids.
- Signs of abuse and addiction should be sought at re-assessment of patients using strong opioids. Routine urine drug testing, pill counts or prescription monitoring should not be used to detect problem use.
- Currently available screening tools should not be relied upon to obtain an accurate prediction of patients at risk of developing problem opioid use before commencing treatment.
- Specialist referral or advice should be considered if there are concerns about rapid-dose escalation with continued unacceptable pain relief, or if >180 mg/day morphine equivalent dose is required.

**Good Practice Points** (based on clinical experience of the guideline development group)

- All patients on strong opioids should be assessed regularly for changes in pain relief, side effects and quality of life, with consideration given to a gradual reduction to the lowest effective dose.
- Opioid rotation should be considered for chronic pain that is likely to respond to opioids, if there are problems with efficacy or side effects.
- There should be careful assessment of pre-existing risk factors for developing opioid misuse. In patients where opioid therapy is indicated, but there is an increased risk of iatrogenic opioid misuse, specialist advice should be sought. The minimal effective dose should be used to avoid increased problems of fracture and overdose that may occur on higher doses.

### **Supplementary Box S2.** Summary of the original SIGN 136 guideline (2013 edition) recommendations for use of gabapentinoids (gabapentin and pregabalin) in the management of adults with chronic non-malignant pain in non-specialist settings

**Recommendations** (based on evidence)

- Gabapentin (titrated up to at least 1,200mg daily) should be considered for the treatment of patients with neuropathic pain
- Pregabalin (titrated up to at least 300mg daily) is recommended for the treatment of patients with neuropathic pain if other first line and second line pharmacological treatments have failed
- Pregabalin (titrated up to at least 300mg daily) is recommended for the treatment of patients with fibromyalgia.
- Flexible dosing may improve tolerability. Failure to respond after an appropriate dose for several weeks should result in trial of a different compound.

### **Supplementary Table S1.** Framework for Enhanced Reporting of Interrupted Time Series Studies (FERITS) Statement

| **Section** | **Item**  **#** | **Recommendation** | **Page #** |
| --- | --- | --- | --- |
| **Title and Abstract** | | | |
| Title and Abstract | 1 | Study design (interrupted time series) indicated in the title or the abstract | **1** |
|  |  | Structured abstract | **2** |
|  |  | Information on target population or study sample | **2** |
| **Introduction** | | | |
| Background | 2 | Scientific background and explanation of rationale | **3** |
| Objectives | 3 | Specific objectives and hypotheses | **4** |
| **Methods** | | | |
| Study Population | 4 | Eligibility criteria for participants, including criteria at different levels in recruitment/sampling plan (e.g., cities, clinics, subjects) | **n/a** |
|  |  | Methods of study population selection (such as codes or algorithms used to identify subjects from routine datasets) | **n/a** |
|  |  | Settings and locations where the data were collected | **5** |
| Study time-period | 5 | Start and end dates of the data included in the study, including reasons for selecting this date range and whether this was the full dataset available or if data was restricted. (Presentation of the full time series as a web appendix recommended) | **5&7** |
|  |  | Time intervals used (e.g. daily, monthly, annual) and the reason for selecting this interval | **6** |
|  |  | Clear definition of the preintervention period, the intervention point (including any transition period) and the post-intervention period | **6&7** |
| Intervention | 6 | Details of the intervention(s) and how and when they were actually administered, specifically including: |  |
|  |  | - Why: Description of the rationale, theory, or goal of the elements essential to the intervention | **6** |
|  |  | - What: Description of what was done including details of any policy changes, procedures, activities and information provided to participants | **6**  **Box S1&S2** |
|  |  | - Who: Who developed, implemented and/or provided the intervention | **6** |
|  |  | - How: Description of the modes of delivery of the intervention and whether it was provided individually, in a group or to a whole population | **6** |
|  |  | - Where: Description of the type(s) of location(s) where the intervention occurred, including any necessary infrastructure or relevant features | **6** |
|  |  | - When: Description of when the intervention was first announced, marketed and delivered, the number of times it was delivered and the duration. Did all groups receive the intervention at the same time | **6** |
|  |  | - Tailoring and modifications: Description of any adaptations or modifications to the intervention during the course of the study | **6** |
|  |  | - Adherence: Report on whether the intervention was compulsory, whether adherence was assessed and any activities to increase compliance or adherence | **6** |
| Outcomes | 7 | Clearly defined outcome measures | **6** |
|  |  | Data source(s) | **5** |
|  |  | Methods used to collect, process, record and extract data; any changes in data collection, processing or recording over time | **5** |
|  |  | Information on validity and reliability of outcome measures | **6&7** |
|  |  | Information on data quality, coverage and completeness over the duration of the study period; any changes in quality, coverage or completeness over time | **6&7** |
| History Bias | 8 | Identification of co-interventions or other concurrent events that might affect the outcome; if no such events exist, clear statement that the intervention was independent of other changes | **6** |
|  |  | Description of design adaptations to mitigate the risk of history bias e.g: adding a control series, using multiple phases or a using a multiple baseline design | **6** |
| Unit of Analysis | 9 | Description of the smallest unit that is being analysed to assess intervention effects (e.g., individual, group, or community) | **7** |
| Statistical Analysis | 10 | Statistical methods used (e.g. segmented Poisson regression, ARIMA etc) | **7** |
|  |  | Appropriateness of a linear model, including description of any tests for linearity and any non-linear terms included | **7** |
|  |  | Detailed description of the a priori impact model and why this was chosen, including allowance for: step or slope change effects, lagged effects, transition phase, floor or ceiling effects. Describe why this model is appropriate for the intervention and outcome under study | **7** |
|  |  | Adjustments for time varying confounders (including seasonality) | **7** |
|  |  | Assessment of autocorrelation and how this was handled | **7&8** |
|  |  | Description of any stratified or subgroup analyses | **7** |
|  |  | Explanation of how missing data were addressed | **n/a** |
|  |  | Discussion of uncertainty in the primary statistical model and description of any additional sensitivity analyses | **n/a** |
|  |  | Statistical software or programs used | **8** |
| **Results** | | | |
| Numbers Analysed | 11 | Report on the number of participants (denominator) included in each analysis for each study condition throughout the study period, particularly when the denominators change for different outcomes. | **9** |
| Population Characteristics | 12 | Description of the baseline demographic and clinical characteristics of the intervention group and any control groups at baseline and throughout the study period (a table is recommended) | **9** |
|  |  | Report on study group equivalence at baseline and statistical methods used to control for baseline differences | **9&10** |
|  |  | Identification of differential changes in population characteristics between study groups throughout the study period and description of statistical methods used to control for differential changes | **n/a** |
| Outcomes and Estimation | 13 | For each primary and secondary outcome, a summary of results for each estimation study condition, and the estimated effect size and a confidence interval to indicate the precision | **9&10** |
|  |  | Report on both relative and absolute changes in the study outcomes following the intervention | **9&10** |
|  |  | Inclusion of null and negative findings | **9&10** |
|  |  | Inclusion of results from testing pre-specified causal pathways through which the intervention was intended to operate, if any | **n/a** |
|  |  | Graphical presentation of the time series for each outcome with the regression line, pre-intervention time period, intervention points and post-intervention period clearly indicated | **Figures 1,2&S1,S2** |
| Ancillary Analyses | 14 | Summary of other analyses performed, including subgroup or restricted analyses and sensitivity analyses, indicating which are pre-specified or exploratory | **10&11** |
| Adverse Events | 15 | Summary of all important adverse events or unintended effects in each study condition (including summary measures, effect size estimates, and confidence intervals) | **n/a** |
| **Discussion** | | | |
| Interpretation | 16 | Interpretation of the results, taking into account study hypotheses, sources of potential bias, imprecision of measures, multiplicative analyses, and other limitations or weaknesses of the study | **12&13** |
|  |  | Discussion of results taking into account the mechanism by which the intervention was intended to work (causal pathways) or alternative mechanisms or explanations | **13** |
|  |  | Discussion of the success of and barriers to implementing the intervention, fidelity of implementation | **13** |
|  |  | Discussion of research, programmatic, or policy implications | **13** |
| Generalizability | 17 | Generalizability (external validity) of the findings, taking into account the study population, the characteristics of the intervention, length of follow-up, incentives, compliance rates, specific sites/settings involved in the study, and other contextual issues | **12** |
| Overall Evidence | 18 | General interpretation of the results in the context of current evidence and current theory | **14** |

### **Supplementary Table S2.** List of opioids (BNF chapter 4.7.2), opioid-containing combination analgesics and gabapentinoid drugs included in the study and the number of items prescribed in Scotland between Q1 2005 and Q2 2020

| **Drug** | **Type** | **Strength** | **Number of prescribed items**  **(% of total)** |
| --- | --- | --- | --- |
| **Opioids** (total: 91,210,542 items) | | | |
| Co-codamol | Combination | Weak | 40,099,832 (44.0) |
| Tramadol hydrochloride | Single | Weak | 14,350,361 (15.7) |
| Co-dydramol | Combination | Weak | 7,430,818 (8.1) |
| Dihydrocodeine | Single | Weak | 7,200,128 (7.9) |
| Methadone | Single | Strong | 7,184,903 (7.9) |
| Morphine | Single | Strong | 5,554,778 (6.1) |
| Oxycodone | Single | Strong | 2,717,417 (3.0) |
| Codeine | Single | Weak | 2,297,259 (2.5) |
| Fentanyl | Single | Strong | 1,361,855 (1.5) |
| Buprenorphine | Single | Strong | 1,215,554 (1.3) |
| Co-codamol with buclizine | Combination | Weak | 536,183 (0.6) |
| Buprenorphine and naloxone | Combination | Strong | 512,987 (0.6) |
| Paracetamol with tramadol | Combination | Weak | 141,102 (0.2) |
| Diamorphine | Single | Strong | 123,883 (0.1) |
| Meptazinol | Single | Weak | 117,240 (0.1) |
| Tapentadol | Single | Strong | 104,494 (0.1) |
| Pethidine | Single | Strong | 100,239 (0.1) |
| Hydromorphone hydrochloride | Single | Strong | 45,658 (0.1) |
| Dipipanone with cyclizine | Combination | Strong | 38,773 (<0.1) |
| Oxycodone and naloxone | Combination | Strong | 36,547 (<0.1) |
| Morphine with cyclizine | Combination | Strong | 17,624 (<0.1) |
| Pentazocine | Single | Strong | 9,986 (<0.1) |
| Co-codaprin | Combination | Weak | 6,370 (<0.1) |
| Ibuprofen with codeine phosphate | Combination | Weak | 5,440 (<0.1) |
| Aspirin with papaveretum | Combination | Strong | 960 (<0.1) |
| Paracetamol, codeine & caffeine | Combination | Weak | 86 (<0.1) |
| Papaveretum | Single | Strong | 64 (<0.1) |
| Pethidine with promethazine | Combination | Strong | 1 (<0.1) |
| **Gabapentinoids** (total: 12,036,499 items) | | | |
| Gabapentin | Single | n/a | 7,529,215 (62.6) |
| Pregabalin | Single | n/a | 4,507,284 (37.4) |

### **Supplementary Table S3.** Mid-year population estimates for Scotland

| **Year** | **Mid-year population estimates (n)** |
| --- | --- |
| **2005** | 5,110,200 |
| **2006** | 5,133,000 |
| **2007** | 5,170,000 |
| **2008** | 5,202,900 |
| **2009** | 5,231,900 |
| **2010** | 5,262,200 |
| **2011** | 5,299,900 |
| **2012** | 5,313,600 |
| **2013** | 5,327,700 |
| **2014** | 5,347,600 |
| **2015** | 5,373,000 |
| **2016** | 5,404,700 |
| **2017** | 5,424,800 |
| **2018** | 5,438,100 |
| **2019** | 5,463,300 |
| **2020** | 5,466,000 |

### **Supplementary Table S4**. Mid-year population estimates for age and gender demographics in Scotland

| **Year** | **Mid-year population estimates (n)** | | | | | |
| --- | --- | --- | --- | --- | --- | --- |
|  | **Female** | **Male** | **0-29 Years** | **30-49 Years** | **50-69 Years** | **70+ Years** |
| **2010** | 2,713,982 | 2,548,218 | 1,884,669 | 1,467,621 | 1,284,671 | 625,239 |
| **2011** | 2,729,600 | 2,570,300 | 1,893,512 | 1,464,320 | 1,310,117 | 631,951 |
| **2012** | 2,736,310 | 2,577,290 | 1,890,570 | 1,450,663 | 1,332,371 | 639,996 |
| **2013** | 2,741,020 | 2,586,680 | 1,887,723 | 1,436,912 | 1,353,549 | 649,516 |
| **2014** | 2,751,070 | 2,596,530 | 1,886,847 | 1,423,205 | 1,374,675 | 662,873 |
| **2015** | 2,762,531 | 2,610,469 | 1,891,211 | 1,413,680 | 1,398,066 | 670,043 |
| **2016** | 2,777,197 | 2,627,503 | 1,897,229 | 1,409,555 | 1,416,588 | 681,328 |
| **2017** | 2,784,500 | 2,640,300 | 1,893,051 | 1,404,236 | 1,420,012 | 707,501 |
| **2018** | 2,789,349 | 2,648,751 | 1,883,269 | 1,401,064 | 1,428,066 | 725,701 |
| **2019** | 2,800,297 | 2,663,003 | 1,877,374 | 1,402,875 | 1,438,350 | 744,701 |
| **2020** | 2,800,788 | 2,665,212 | 1,860,869 | 1,404,025 | 1,445,459 | 755,647 |

### **Supplementary Table S5.** Results of the controlled interrupted time series analysis of weak^1^ compared to strong opioid^2^ prescribing rates in Scotland

|  | **Estimate**  **(95% Confidence Interval)** | **Standard Error** | **P value** |
| --- | --- | --- | --- |
| Difference in Intercept, β_4_ | 154.12 (150.22, 158.02) | 1.97 | <0.01 |
| Difference in Pre-Intervention Trend, β_5_ | 0.54 (0.36, 0.73) | 0.09 | <0.01 |
| Difference in Change in Level, β_6_ | 0.41 (-5.54, 6.36) | 3.00 | 0.89 |
| Difference in Change in Trend, β_7_ | -1.72 (-2.07, -1.37) | 0.18 | <0.01 |
| Difference in Post-Intervention Trend, β_5+7_ | -1.18 (-1.45, -0.91) | 0.13 | <0.01 |
| Durbin-Watson Statistic | 1.64 | n/a | n/a |

β_4-7_ are coefficients from the controlled interrupted time series analysis model; the intercept represents the outcome at the start of the study period.

^1^Weak opioids are the following and their compounds: codeine, dihydrocodeine, meptazinol and tramadol

^2^Strong opioids are the following and their compounds: buprenorphine, diamorphine, dipipanone, fentanyl, hydromorphone, methadone, morphine, oxycodone, papaveretum, pentazocine, pethidine and tapentadol

### **Supplementary Table S6.** Results of the single interrupted time series analyses of female and male opioid prescribing rates in Scotland

|  | **Female** | | | **Male** | | |
| --- | --- | --- | --- | --- | --- | --- |
|  | **Estimate**  **(95% Confidence Interval)** | **Standard Error** | **P value** | **Estimate**  **(95% Confidence Interval)** | **Standard Error** | **P value** |
| Intercept, β_0_ | 291.76 (284.16, 299.36) | 3.75 | <0.01 | 195.03 (189.77, 200.28) | 2.59 | <0.01 |
| Pre-Intervention Trend, β_1_ | 2.73 (1.94, 3.51) | 0.39 | <0.01 | 2.09 (1.54, 2.63) | 0.27 | <0.01 |
| Change in Level, β_2_ | 7.15 (-1.90, 16.21) | 4.47 | 0.12 | 4.73 (-1.54, 10.99) | 3.09 | 0.14 |
| **Change in Trend, β_3_** | **-3.26 (-4.13, -2.39)** | **0.43** | **<0.01** | **-2.52 (-3.13, -1.92)** | **0.30** | **<0.01** |
| **Post-Intervention Trend, β_1+3_** | **-0.54 (-0.94, -0.13** | **0.20** | **0.01** | **-0.44 (-0.71, -0.16)** | **0.13** | **<0.01** |
| **Relative Change, %^a^** | **-19.11 (-23.61, -13.13)** | **n/a** | **n/a** | **-21.54 (-25.88, -15.64)** | **n/a** | **n/a** |
| Durbin-Watson Statistic | 2.27 | n/a | n/a | 2.16 | n/a | n/a |

β_0-3_ are coefficients from the single-group interrupted time series analysis model; the intercept represents the outcome at the start of the study period; the relative change is calculated compared to the predicted value at the same time point had the pre-intervention trend continued.

^a^Calculated at quarter 2 202095% confidence interval calculated using model bootstrapping

### **Supplementary Table S7.** Results of the controlled interrupted time series analysis of females compared to male opioid prescribing rates in Scotland

|  | **Estimate**  **(95% Confidence Interval)** | **Standard Error** | **P value** |
| --- | --- | --- | --- |
| Difference in Intercept, β_4_ | 96.73 (87.65, 105.82) | 4.56 | <0.01 |
| Difference in Pre-Intervention Trend, β_5_ | 0.64 (-0.30, 1.58) | 0.47 | 0.18 |
| Difference in Change in Level, β_6_ | 2.43 (-8.41, 13.26) | 5.44 | 0.66 |
| Difference in Change in Trend, β_7_ | -0.74 (-1.78, 0.31) | 0.52 | 0.16 |
| Difference in Post-Intervention Trend, β_5+7_ | -0.10 (-0.58, 0.38) | 0.24 | 0.68 |
| Durbin-Watson Statistic | 2.25 | n/a | n/a |

β_4-7_ are coefficients from the controlled interrupted time series analysis model; the intercept represents the outcome at the start of the study period.

### **Supplementary Table S8.** Results of the single interrupted time series analyses of opioid prescribing rates by age category in Scotland

|  | **Estimate**  **(95% Confidence Interval)** | **Standard Error** | **P value** |
| --- | --- | --- | --- |
| **0-29 years** | | | |
| Intercept, β_0_ | 35.56 (33.34, 37.78) | 1.10 | <0.01 |
| Pre-Intervention Trend, β_1_ | 0.33 (0.10, 0.56) | 0.11 | 0.01 |
| Change in Level, β_2_ | 1.47 (-1.18, 4.12) | 1.31 | 0.27 |
| Change in Trend, β_3_ | -0.74 (-1.00, -0.49) | 0.13 | <0.01 |
| Post-Intervention Trend, β_1+3_ | -0.41 (-0.53, -0.30) | 0.06 | <0.01 |
| Relative Intervention Effect, %^a^ | -36.13 (-44.43, -22.56) | n/a | n/a |
| Durbin-Watson Statistic | 1.80 | n/a | n/a |
| **30-49 years** | | | |
| Intercept, β_0_ | 235.12 (226.01, 244.22) | 4.50 | <0.01 |
| Pre-Intervention Trend, β_1_ | 4.33 (3.39, 5.27) | 0.47 | <0.01 |
| Change in Level, β_2_ | 10.42 (-0.44, 21.27) | 5.36 | 0.06 |
| Change in Trend, β_3_ | -5.47 (-6.52, -4.43) | 0.52 | <0.01 |
| Post-Intervention Trend, β_1+3_ | -1.15 (-1.67, -0.62) | 0.26 | <0.01 |
| Relative Intervention Effect, %^a^ | -31.64 (-35.43, -26.81) | n/a | n/a |
| Durbin-Watson Statistic | 1.62 | n/a | n/a |
| **50-69 years** | | | |
| Intercept, β_0_ | 393.82 (382.24, 405.40) | 5.72 | <0.01 |
| Pre-Intervention Trend, β_1_ | 2.94 (1.74, 4.14) | 0.59 | <0.01 |
| Change in Level, β_2_ | 4.30 (-9.51, 18.11) | 6.82 | 0.53 |
| Change in Trend, β_3_ | -3.21 (-4.54, -1.88) | 0.66 | <0.01 |
| Post-Intervention Trend, β_1+3_ | -0.27 (-0.83, 0.30) | 0.27 | 0.34 |
| Relative Intervention Effect, %^a^ | -15.29 (-21.33, -6.24) | n/a | n/a |
| Durbin-Watson Statistic | 2.43 | n/a | n/a |
| **70+** | | | |
| Intercept, β_0_ | 599.15 (589.01, 609.28) | 5.01 | <0.01 |
| Pre-Intervention Trend, β_1_ | 0.81 (-0.24, 1.86) | 0.52 | 0.12 |
| Change in Level, β_2_ | 13.61 (1.53, 25.70) | 5.97 | 0.03 |
| Change in Trend, β_3_ | -3.46 (-4.62, -2.29) | 0.57 | <0.01 |
| Post-Intervention Trend, β_1+3_ | -2.64 (-3.18, -2.11) | 0.26 | <0.01 |
| Relative Intervention Effect, %^a^ | -12.04 (-16.25, -6.33) | n/a | n/a |
| Durbin-Watson Statistic | 2.47 | n/a | n/a |

β_0-3_ are coefficients from the single-group interrupted time series analysis model; the intercept represents the outcome at the start of the study period; the relative change is calculated compared to the predicted value at the same time point had the pre-intervention trend continued.

^a^Calculated at quarter 2 2020. 95% confidence interval calculated using model bootstrapping

### **Supplementary Figure S1.** Opioid prescribing time series across Scotland by gender, before and after the publication of SIGN 136 in December 2013 (red dashed line). The solid lines represent the prescribing trend derived from the interrupted time series analysis.

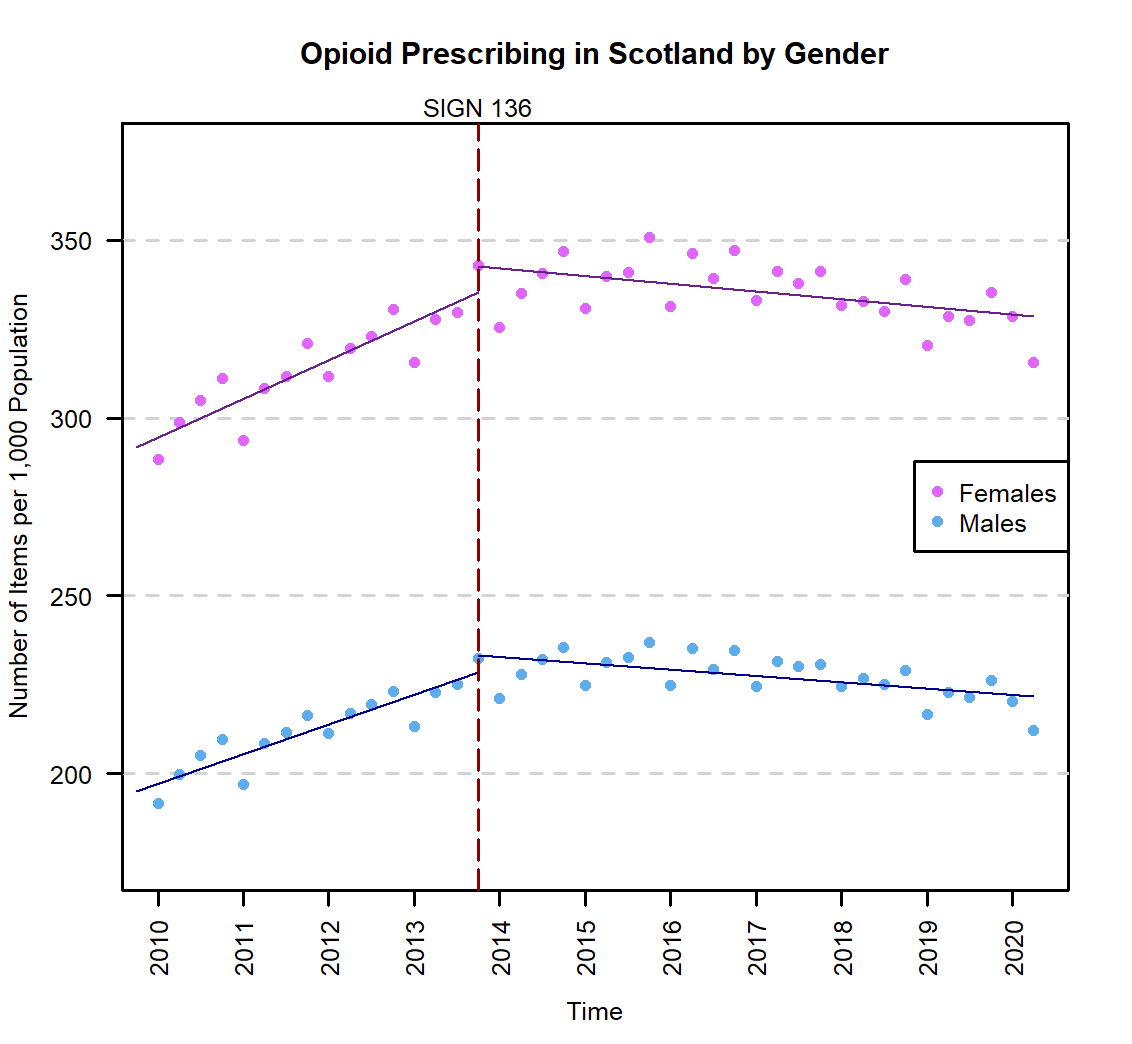

### **Supplementary Figure S2.** Opioid prescribing time series across Scotland by age, before and after the publication of SIGN 136 in December 2013 (red dashed line). The solid lines represent the prescribing trend derived from the interrupted time series analysis.

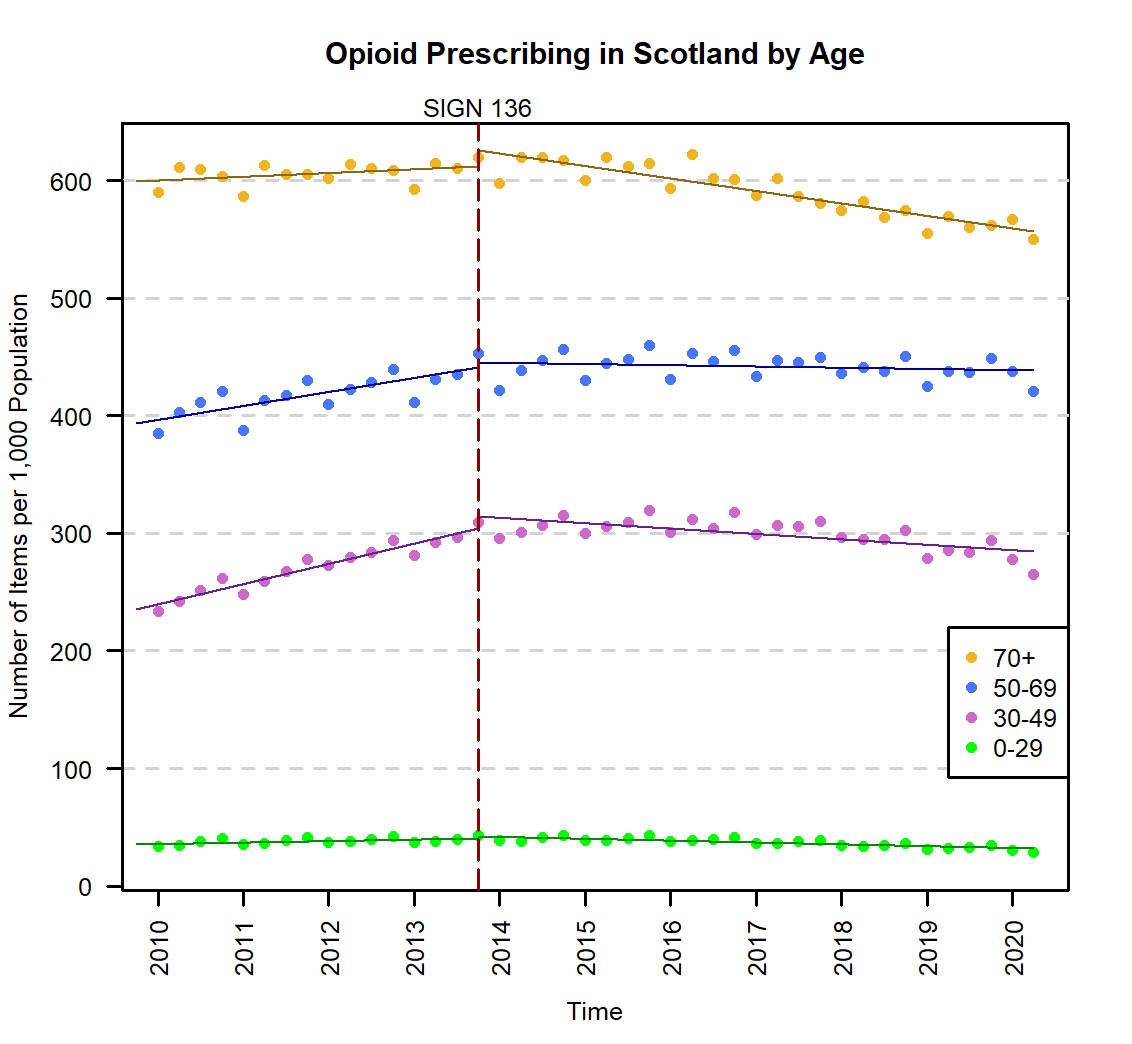
